# Modelling the effect of the interaction between vaccination and non-pharmaceutical measures on COVID-19 incidence

**DOI:** 10.1101/2021.11.29.21266986

**Authors:** Atsegine Canga, Gorka Bidegain

## Abstract

Since December 2019, the novel severe acute respiratory syndrome coronavirus 2 (SARS-CoV-2) has spread rapidly from Wuhan (China) across the globe, affecting more than 200 countries by mid-2021, with over 190 M reported cases and around 4 M fatalities. During the first year of the pandemic, affected countries implemented a variety of non-pharmaceutical interventions to control virus transmission. In December 2020, countries started administering several authorised vaccines under a limited supply scenario. In this context, the aim of this study was to develop a SEIR-type continuous-time deterministic disease model, to determine the impact of interaction between different vaccination scenarios and levels of protection measures on disease incidence. For this, the model incorporates (i) a protection measure including low (self-protection), medium (mobility limitation), high (closure of indoor facilities) and very high (lockdown) protection levels, (ii) quarantine for confirmed cases, and (iii) vaccination rate and efficacy of four type of vaccines (Pfizer, Moderna, Astra Zeneca or Janssen). The model was verified and evaluated using the response timeline and vaccination strategies and rates in the Basque Country (N. Spain). Once the model performance was validated, different initial phase (when 30% of the population is vaccinated) vaccination scenarios were simulated, including (i) a realistic vaccine limited supply scenario, and (ii) four potential full vaccine supply scenarios where a unique vaccine type is administered. Some differences in disease incidence were found between vaccination scenarios for low and medium-level protection measures. However, regardless of the administered vaccine, a high-level protection scenario is the most effective to control the virus transmission and disease mortality in the studied initial phase of vaccination. The results obtained here may vary in further studies since there may be some unpredictable factors/covariates. With this in mind, the model here could be easily applied to other regions or countries, modifying the strategies implemented and initial conditions.

## 1 Introduction

The novel severe acute respiratory syndrome coronavirus 2 (SARS-CoV-2) or coronavirus disease 2019 (COVID-19), was first detected in Wuhan, China, in December 2019, as the cause of a pneumonia of unknown aetiology (Boni et al., 2020; Li et al., 2020; Mwalili et al., 2020). The SARS-CoV-2 rapidly spread all over the globe and on March 11, 2020, the World Health Organization (WHO) declared COVID-19 a global pandemic (Cucinotta & Vanelli, 2020; Mwalili et al., 2020; Rothan & Byrareddy, 2020). After more than a year and a half since the pandemic was declared, according to the WHO, there are 200 M cases reported and 4 M deaths worldwide (ECDC, 2021).

The SARS-CoV-2 transmission is through exposure by (i) inhalation of very fine respiratory droplets and aerosol particles released by infected individuals, mostly between people at close range (Tang et al., 2021) (ii) deposition of respiratory droplets and particles on exposed mucous membranes (mouth, nose, or eyes) by direct splashes and sprays, and (iii) touching mucous membranes with hands that have been soiled by touching surfaces with virus (Liu et al., 2020; Sheng, 2020; WHO, 2021a). The virus transmission in indoor settings has been the main transmission pathway when ventilation is not sufficient (Atalan, 2020; Baghat et al., 2020; Jayaweera et al., 2020; WHO, 2021a).) The main outbreaks have been related to explosive super events in indoor settings or facilities such as family gatherings, long-term health facilities, restaurants, bars and clubs (e.g. Chau et al., 2021), being these principal responsible of the dynamics and shape of the COVID-19 transmission (Althouse et al., 2020).

Due to the rapid spread of the virus through these events and the lack of effective pharmaceutical treatments for the disease, particularly at the beginning of the pandemic, important control measures have been implemented worldwide: quarantine of people suspected of being exposed to COVID-19, isolation/quarantine of confirmed cases, use of face masks in public, contact tracing, social distancing, closing of indoor settings (public spaces, restaurants, bars, etc.) and schools and universities, working from home, confinement of regions with a high incidence of the virus, to total lockdown of the country to slow down the COVID-19 outbreak (Atalan, 2020; CDC (2022); ECDC, 2020; Iboi et al., 2020; MacIntyre et al., 2020; WHO, 2020).

In addition to these measures, since the pandemic began, the governments of disease-impacted countries have been working on the development and implementation of strategies to return to “normal life”, including the development of several vaccines against COVID-19 (CDC, 2021; WHO, 2022). To date, a variety of vaccines have been approved by the European Medicine Agency (EMA), under a conditional marketing authorisation due to the emergency situation, and many others are under development. These vaccines have different efficacy, understood as the percentage reduction in disease incidence, based on clinical trials (EMA, 2021). Vaccination is based on the fact that if a fraction of the population is immune to that pathogen, the susceptible host numbers decrease, so the impact of infected individuals is limited (Randolph & Barreiro, 2020; Sariol & Perlman, 2020). Herd immunity originates when a sufficiently large proportion of the population is immune to the disease (Omer et al., 2020; Randolph & Barreiro, 2020). The percentage of the population that needs to be vaccinated to achieve herd immunity varies with each disease. Several studies have concluded that to achieve COVID-19 herd immunity and relax protection measures around 50% to 70% of the population should be vaccinated (Clemente-Suárez et al., 2020; Kim et al., 2021). The immediate goal of the global COVID-19 vaccination strategy is to minimize deaths, severe disease incidence and reduce the risk of new variants. This requires fully vaccinating at least 70% of the world’s population, accounting for most adults and adolescents and for the vast majority of those at risk of serious disease (WHO, 2021b). Consequently, initial vaccination phases such as the one studied here (around 30% of the population vaccinated) may require the maintenance of some level of non-pharmaceutical protection measures to control disease transmission.

In this context, the modelling approach is a determinant tool to analyse COVID-19 disease dynamics and support the development of public health policies (Wong et al., 2021). Most models for the COVID-19 pandemic are single-population continuous compartmental SEIR Kermack-McKendrick-type models, constructed using ordinary differential equation (ODE) systems (Guirao et al., 2020; Li et al., 2020; Tang, Bragazzi, et al., 2020; Wu et al., 2020). Compartmental models are a very common infectious modelling approach where the population is assigned to compartments with labels (S, Susceptible; E, Exposed; I, Infectious; R, Recovered) and individuals may progress between compartments. For COVID-19 models apart from the S and I compartments, E and R compartment are essential since: (i) there is a significant latency period during which individuals have been infected but are not yet infectious themselves, so they are exposed, and (ii) sick individuals recovered from disease they are not infectious (I) and they are immune for some months so they cannot be considered susceptible.”

These population disease models may be basic in order to capture certain disease dynamic complexities. However, for any emerging pandemic, they are essential, first to develop the theoretical basis for the understanding of pathogen transmission processes and mechanisms, and second, to explore disease spread control measures. A limitation when modelling COVID-19 transmission is that only confirmed cases are known. There is a fraction of non-reported positive cases, ranging between 10-70% of the total, that correspond to people that do not get tested or are asymptomatic to the disease. Hence, models such us the ones developed by the Imperial College of London, estimate that this fraction will show a higher number of cases compared to the reported data (Giattino, 2020). Given the uncertainty surrounding the situation after COVID-19 vaccination programmes, models estimating these unconfirmed cases, such as the one presented here, can be particularly useful for exploring different scenarios of immunisation through the vaccination effect on disease spread limitation.

This work is focused on the development of a deterministic SEIR transmission model to analyse the impact of the interaction between different vaccination scenarios, regarding vaccinaton rate and efficacy, and different levels of non-pharmaceutical protection measures (from the use of mask to lockdown) on disease incidence and mortality. The model scenarios are set for the initial phase of the COVID-19 vaccination (i.e. when around 30% of the population is vaccinated) and evaluated on the response timeline of the first and second waves of the pandemic in the Basque Country (N Spain), one of the regions reporting highest disease incidence in Europe.

## 2 Methods

### 2.1 Model description and mathematical theory

The model here is an extension of a Kermack-McKendrick-type model (Kermack & McKendrick, 1927). It is a deterministic SEIR transmission model that accounts for important characteristic for understanding COVID-19 disease dynamics, such as (i) incubation period, (ii) a protection measure ranging from low-level protection (self-protection; use of a mask, hygiene and social distancing), medium-level protection (mobility limitation), high-level protection (adding indoor facilities closure) and very high-level protection (lockdown), (iii) quarantine for confirmed cases, and (iv) vaccination rate and efficacy.

The model is a one-population compartmental model, continuous in time, unstructured in spatial or age terms, and configured to simulate the dynamics of COVID-19 transmission processes caused by susceptible individuals contacting infected individuals or environments with infectious particles released by infected individuals. The compartmental models to describe pathogen transmission are the most frequently used class of models in epidemiology (Diekmann & Heesterbeek, 2000). Individuals can take on a finite number of discrete states, and each state is representative of a subpopulation of individuals at a given time (Table 1). These compartments and states, in consequence, are defined as the variables of the model. These variables together with the associated parameters satisfy a system of ODEs describing the dynamics of the host-pathogen system.

**Table 1.** Variable description with model assumptions and initial values obtained by model validation assuming confirmed cumulative mortality data. Initial values in this table are the ones used in the model verification/evaluation on the first wave data. For the verification/evaluation on the second wave data and simulations of vaccination scenarios, changes to these initial values are defined further on.

| **Variable** | **Description and modelling assumptions** | **Initial conditions**<br>**(individuals)** |
| --- | --- | --- |
| *N* | Population in the Basque Country | 2199711 |
| *S* | Population of non-quarantined susceptible individuals | 2199671 |
| *E* | Population of susceptible exposed individuals; infected individuals with no symptoms and no infectivity | 30 |
| *I* | Population of infected individuals with infective capacity and not quarantined; asymptomatic or not reported cases | 10 |
| *Q* | Population of quarantined infected individuals; reported cases | 0 |
| *R* | Patients recovered from COVID-19 | 0 |
| *D* | Individuals deceased due to COVID-19 | 0 |
| *V* | Vaccinated and protected population against COVID-19 | 0 |

The model here includes seven compartments (i.e. variables or subpopulations) (Table 1) and each state assumes the following: (1) *S* stands for susceptible subpopulation that can become exposed to the virus by contact with an infected individual or with infectious particles released by an infected individual; (2) *E* represents the population exposed to the virus after being in contact with an infected individual or infected environment; (3) *I* represents the infected subpopulation with individuals coming from the exposed subpopulation after the corresponding incubation period of the virus (five days on average (Lauer et al. 2020; Rǎdulescu et al., 2020)); The *I* subpopulation is assumed to represent asymptomatic cases, non-confirmed and non-isolated symptomatic cases, and cases that are not yet or are not quarantined; consequently this subpopulation can be considered the source of the infection in the model. According to the WHO, although asymptomatic people can spread the virus (Rǎdulescu et al., 2020), they are most infectious in the early stages of a symptomatic stage, so that the majority of infections are caused by symptomatic individuals (WHO, 2020), (4) a fraction of this *I* subpopulation is the pool of infected individuals, represent confirmed and quarantined people, representing the *Q* subpopulation. This subpopulation of quarantined people after being diagnosed with COVID-19 includes home isolated and hospitalised patients; (5) *R* represents the population that has recovered from the disease and is immune to disease during a certain period of immunisation time; (6) *V* subpopulation represents vaccinated individuals with protection against the virus and (7) *D* represents individuals that die due to COVID-19, that is, this variable tracks cumulative deaths (Table 1).

The variables or subpopulations of the host population are defined with respect to the number of individuals in the studied territory. Thus, the initial population *N* for the model is 2199711 individuals based on demographic data of the Basque Country Institute of Statistics (BIS) (BIS, 2020). The model specifies an open population where birth of new susceptible individuals is a function of the total population N. Since this is a novel coronavirus, initially everyone is susceptible to COVID-19. The model assumes some individuals were already exposed and infected at simulation day 1 (March 1) (Table 1). These values are obtained by model fitting against real cumulative mortality data (BHD, 2020) and considering that at least 33% of the cases are asymptomatic, not confirmed and able to infect, but not under quarantine (Pollán et al., 2020).

Another feature of the COVID-19 virus, is the incubation period, which is relatively long and an individual is able to infect others before being diagnosed (Rǎdulescu et al., 2020). In general, some model features and specific assumptions such as protection measures defined by the parameters in Table 2 may result in some predictive limitations (see section 2.7). Based on all these assumptions and simplifications, the basic model for the transmission dynamics of COVID-19 is given by the following deterministic system of nonlinear differential equations:

**Table 2.** Baseline model parameters with a brief description and default values used for the model.

| Parameter | Description | Value | Range | Units | References |
| --- | --- | --- | --- | --- | --- |
| $\gamma$ | Incubation rate | $2.0 \times 10^{-1}$ | 0.1 - 0.3 | day<sup>-1</sup> | Amira et al. (2020)<br>Dhouib et al. (2021)<br>Lauer et al. (2020)<br>Rădulescu et al. (2020) |
| $r_1$ | Recovery rate for I | $7.0 \times 10^{-2}$ | 0.5 - $1 \times 10^{-1}$ | day<sup>-1</sup> | Rădulescu et al. (2020)<br>Yang et al. (2021) |
| $r_2$ | Recovery rate for Q | $5.0 \times 10^{-2}$ | | | |
| $q$ | Quarantine rate | $6.7 \times 10^{-1}$ | 0.5 - 0.8 | day<sup>-1</sup> | Tang, Xia, et al. (2020).<br>Yang et al. (2021) |
| $\beta$ | Infection rate | 1.1 | 0.6 - 1.7 | day<sup>-1</sup> | Lin et al. (2020) |
| $b$ | Birth rate | $2.7 \times 10^{-5}$ | | | EUSTAT (2022a) |
| $d$ | Disease mortality rate | $5.7 \times 10^{-3}$ | 0.1 - $0.6 \times 10^{-1}$ | day<sup>-1</sup> | López & Rodó, (2021)<br>Rădulescu et al., (2020) |
| $m$ | Natural mortality rate | $2.7 \times 10^{-5}$ | | day<sup>-1</sup> | EUSTAT (2022b) |
| $\nu$ | Vaccine protection rates (Pfizer, Moderna, Astra Zeneca, Janssen) | $1.3 \times 10^{-3}$ | | day<sup>-1</sup> | Estimated in this study (Table 3) |
| | | $1.8 \times 10^{-4}$ | | | |
| | | $3.1 \times 10^{-5}$ | | | |
| | | $9.8 \times 10^{-5}$ | | | |
| $\alpha$ | Protection rate:<br>Low (mask use, hygiene, social distancing), medium (mobility restrictions), high (mobility and indoors closure) very high (lockdown) | $5.0 \times 10^{-3}$ | 0.3 - $3 \times 10^{-2}$ | day<sup>-1</sup> | López & Rodó, (2021)<br>Estimated in this study |
| | | $7.5 \times 10^{-3}$ | | | |
| | | $10 \times 10^{-3}$ | | | |
| | | $25 \times 10^{-3}$ | | | |
| $\sigma$ | Immunity loss rate | $1.1 \times 10^{-2}$ | | day<sup>-1</sup> | Dan et al.(2021) |

### 2.2 Model Equations

The subpopulations of the model satisfy a system of ODEs describing the dynamics of the host-virus association. Variables and parameters of these equations are described in Tables 1 and 2, respectively. The numerical model for this ODE system is programmed in Matlab R2018a. The set of coupled differential equations is solved with a fourth–order predictor corrector scheme, using the Adams Bashforth predictor and the Adams-Moulton corrector. The differential equation system comprises the following differential equations:

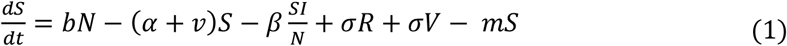

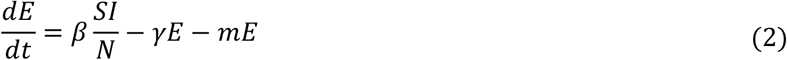

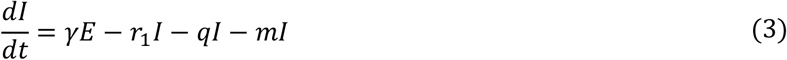

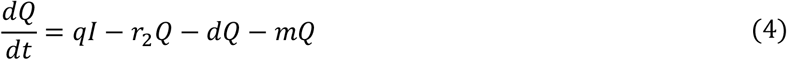

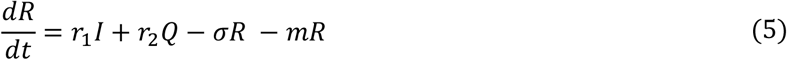

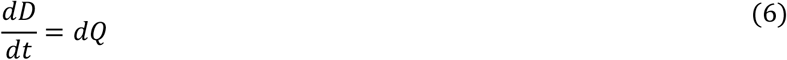

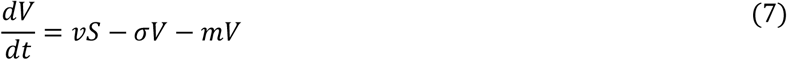

*Equation (1):* The change in the number of susceptible individuals *S*, is a balance between (i) the loss of individuals due to protection measures (use of mask, social distancing, mobility restrictions, indoor settings closure, lockdown) and vaccination, virus transmission and background mortality and (ii) the gain of individuals from births, and recovered and vaccinated individuals who lost immunisation.

*Equation (2):* The change in the number of exposed individuals *E*, is a balance between (i) the gain of individuals due to virus transmission, and the (ii) loss of individuals because of background mortality or the end of the incubation period in exposed individuals.

*Equation (3):* The change in the number of infected individuals *I*, is a balance between the (ii) gain of individuals due to the end of incubation period in exposed individuals, and the (ii) loss of individuals because of background mortality, recovered individuals and the subtraction of the proportion of isolated/quarantined individuals.

*Equation (4):* The change in the number of quarantined individuals *Q*, is a balance between (ii) the gain of individuals due to the proportion of the infected subpopulation who are symptomatic and theoretically recorded as confirmed cases and quarantined individuals, and (ii) the loss of individuals because of disease mortality, recovered individuals and background mortality.

*Equation (5):* The change in the number of recovered individuals *R*, is a balance between the gain of recovered individuals from infection including quarantined individuals, and the loss of recovered individuals who lost immunisation and background mortality.

*Equation (6):* The change in the number of deceased infected individuals *D* (cumulative mortality), is represented by the gain of individuals due to disease mortality from quarantined (and eventually hospitalised) cases.

*Equation (7):* The change in the number of vaccinated and protected individuals *V* against COVID-19, is a balance between the gain of individuals due to immunisation through vaccination, and the loss of individuals due to the loss of immunisation, after a certain period of time, and background mortality.

### 2.3 Data

#### Epidemiological data

Data on infection-confirmed cases, cumulative deaths and vaccination in the Basque Country, were obtained from the open data system on the BHD website (BHD, 2020). The model uses realistic initial conditions for the variables and keeps parameters in the range of recent research findings (see Table 2). The new daily confirmed cases and cumulative mortality data were used for comparison with modelling results and for model verification and evaluation.

#### Vaccination data

The first mass vaccination programme in the Basque Country started on December 27, 2020, reporting by then 117000 cases and 3030 fatalities (BHD, 2020). Since then to June 8, 2021, four vaccines were administered in the Basque Country: Pfizer, Moderna, Astra Zeneca and Janssen. For two-dose vaccines, the time interval between doses is three weeks for Pfizer, four weeks for Moderna and 12 weeks for Astra Zeneca. The model uses the initially reported vaccine efficacy values (EMA, 2021) in Table 3 and 4. This efficacy values are reached after seven days from the booster dose for Pfizer and after 14 days for the Moderna and Astra Zeneca vaccines. For the single dose Janssen vaccine, the reported efficacy is achieved after 14 days. As the vaccine efficacy is very labile and dynamic depending in many factors such as virus variants, updated data on vaccine efficacy for further modelling studies can be obtained in product reports of vaccines in EMA (2022)

**Table 3.** Estimated vaccination and protection rates using the Basque Countries’ vaccination data (BHD, 2020) and the initial efficacy rates reported from EMA (EMA, 2021).

| Vaccine type | Complete doses (CD) (individuals) | Vaccination rate (VR) (individuals/day) | Vaccine efficacy (VE) | Vaccination protection rate (VPR) (model) |
| --- | --- | --- | --- | --- |
| **Pfizer/BionTech** | 496894 | 3105 | 94.6% | $1.3 \cdot 10^{-3}$ |
| **Moderna** | 72164 | 451 | 93.6% | $1.8 \cdot 10^{-4}$ |
| **Astra Zeneca** | 19079 | 119 | 60.0% | $3.1 \cdot 10^{-5}$ |
| **Janssen** | 55841 | 349 | 67.0% | $9.8 \cdot 10^{-5}$ |

**Table 4.**
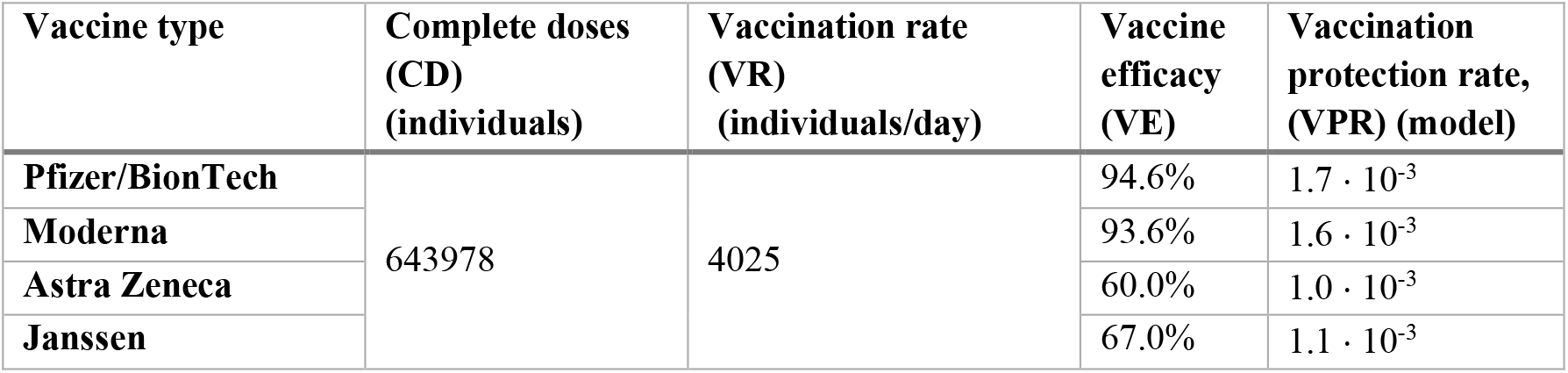
Estimated vaccination and protection rates for unique vaccine administration scenarios and the efficacy reported from EMA (EMA, 2021).

Since the start of the vaccination programme on the 27^th^ of December, up until the 8^th^ of June, 643,978 complete doses of COVID-19 vaccines were administered, distributed by vaccine type as in Table 3. This administration strategy, due to vaccine supply limitations in the Basque Country, envisaged the administration of four vaccines with the following percentages in terms of population: 77.2% Pfizer, 11.2% Moderna, 3.0 % Astra Zeneca and 8.7% Janssen (BHD, 2020).

The average number of vaccines administered per day (vaccination rate, VR), was estimated from data obtained from the vaccination bulletin of the BHD (BHD, 2020). According to the number of complete doses administered and the efficacy of each vaccine, a vaccine-specific vaccination protection rate was estimated for the model (Table 3) as follows:

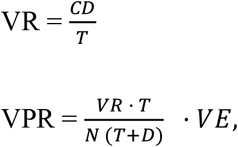

where T is 164 days, from December 27 to June 8, and D is the number of days needed after the second dose was administered for the vaccine to achieve peak efficacy. N is the total population in the Basque Country (N=2199711).

### 2.4 Descriptive analysis of the epidemic curve for the Basque Country

The change in the number of daily COVID-19 new cases in the Basque Country over time, from March 1, 2020 to June 8, 2021 is described in terms of the non-pharmaceutical interventions adopted by the BHD (Figure 1). Results including simulations for model verification/validation and simulations for vaccination scenarios were discussed in terms of this initial descriptive analysis

**Figure 1.**
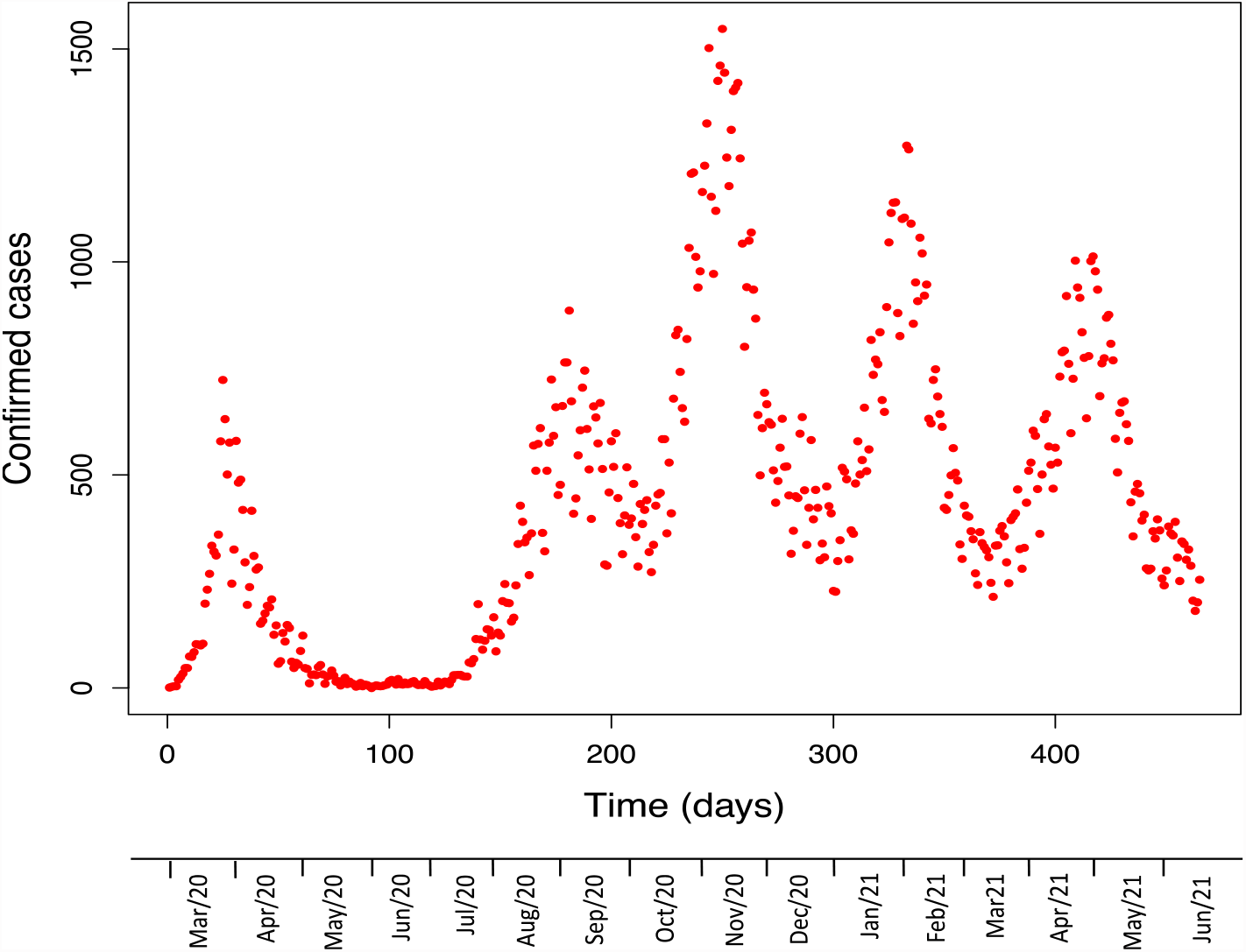
Confirmed cases in the Basque Country from March 1, 2020, to June 8, 2021 (BHD, 2020).

### 2.5 Model verification and evaluation

The model was verified and evaluated with simulations for the first and second waves of the pandemic in the Basque Country using the parameter values in Table 2. Model verification consisted of showing that the simulation model was correct, complete, and coherent. This required analysing (1) static tests involving a structured examination of the formulas, algorithms, and codes used to implement the model, by several reviewers experts in population modelling (see acknowledgements section), and (2) dynamic tests, where the computer program was run under different conditions of disease mortality and quarantine rate. This analysis were critical to ensure that results produced were correct, according to the conceptual model, and consistent with expectations of reviewers. Model evaluation was carried out against confirmed cases and cumulative mortality. Simulations and descriptions of model verification and evaluation are given in the results section.

### 2.6 Modelling vaccination scenarios

Considering the degree of protection of each vaccine in terms of the estimated vaccination protection rate together with different nonpharmaceutical interventios (low, medium and high protection levels (Table 1) the effects of five different vaccination strategies were tested within the model.

#### Realistic scenario with limitations on vaccine supply

In the first scenario, the model tested the effect on infected cases, based on the vaccination strategy followed in the Basque Country. This realistic limited vaccine supply scenario envisages the administration of four vaccines, with the percentages in terms of vaccinated population estimated from Table 3. The model uses vaccine protection rates estimated in Table 3.

#### Realistic scenarios with full vaccine supply

Four full vaccine supply scenarios were simulated where a unique vaccine type administration is possible. For these simulations, the model uses new vaccination protection rates estimated in Table 4, as in *Vaccination data* section, considering that the population receives a unique vaccine.

### 2.7 Model limitations

The four nonpharmaceutical protection levels (Table 2) are considered fully and successfully applied. However, this is a simplification of reality and may result in an underestimation of disease incidence; since violation of low-level protection measures such as the use of mask or social distancing may be much higher than strong measured such as lockdown.

For such a rapid spread of COVID-19 the population is assumed to be constant in terms of the difference between births and natural deaths, i.e. the birth rate is assumed to be the same as the natural death rate (Table 2). In the case of the studied region the birth rate is slightly lower than the mortality rate (EUSTAT, 2022a, b), however this assumption is overestimating the S population.

A seroprevalence study carried out in Spain estimated 33% of the cases as asymptomatic (Pollán et al., 2020). The quarantine rate in the model (0.67 day ^-1^, see Table 2) was estimated based on this study result and assuming that asymptomatic are non-confirmed cases and hence, non-quarantined people. This assumption could be underestimating the quarantined population particularly beyond third wave when testing was common to close contacts of Covid-19.

When modelling realistic vaccine scenarios with vaccine full supply caution is required to interpret the results (see section 3.3) quantitatively; since the model is dealing with multiple dimensions, latent covariates, a local region without %100 certain data and four vaccine types.

### 2.8 Descriptive analysis of the curve of infection cases

The curve of new daily confirmed cases by the BHD (BHD, 2020) (Figure 1) is described below in terms of the non-pharmaceutical interventions’ chronological sequence. This detailed analysis is essential for the selection of simulation cases with different protection measures and later discussion.

The first epidemic wave in the Basque Country in terms of confirmed cases started with the first case of COVID-19 on February 28, 2020. This **first wave** reached its peak on the March 24, with 723 confirmed new cases (Figure 1). Previously, on March 13, schools were closed in all regions of Spain including the Basque Country and a nationwide lockdown (confinement) was declared, banning all public events.

As the confirmed cases were decreasing, on April 13 the partial lifting of the lockdown started, allowing people to go out by age groups at certain times during the day. On May 4, the lockdown was totally lifted, starting as a phase 0, called the “new normality”. Mobility all around the country was permitted in summer and indoor public facilities were open. On July 8, mask use in public areas became mandatory for people older than six years old. By mid-July, daily cases started to increase consistently, and the **second wave** reached its peak on August 28, with 886 confirmed new cases (Figure 1).

The peak of the **third wave** reached on November 5, with 1547 new confirmed cases. When cases started to increase on October 27, the activity and mobility was limited to daytime, and it was prohibited from 11pm to 6am. Also, the region of the Basque Country was closed and travel between regions was forbidden. Only groups with a maximum of six people were allowed. And on November 7, bars and restaurants were closed until December 12, likewise limiting capacity at indoor places.

A **fourth wave** peak was recorded on January 27, 2021, with 1274 new cases. The **fifth wave** started in mid-March, reaching its peak on April 21, with a maximum of 1013 new cases.

### 2.9 Model verification and evaluation

From the set of simulation results obtained during model verification and evaluation, eight simulation cases for the first two waves of the pandemic in the Basque Country are described here: Cases 1-4 (Figure 2) were simulated for the first wave and Cases 5-8 for the second wave (Figure 3). The simulations were run with the initial conditions described in Table 1 and parameter values described in Table 2. Changes to these values for specific simulations are described for each case.

**Figure 2.**
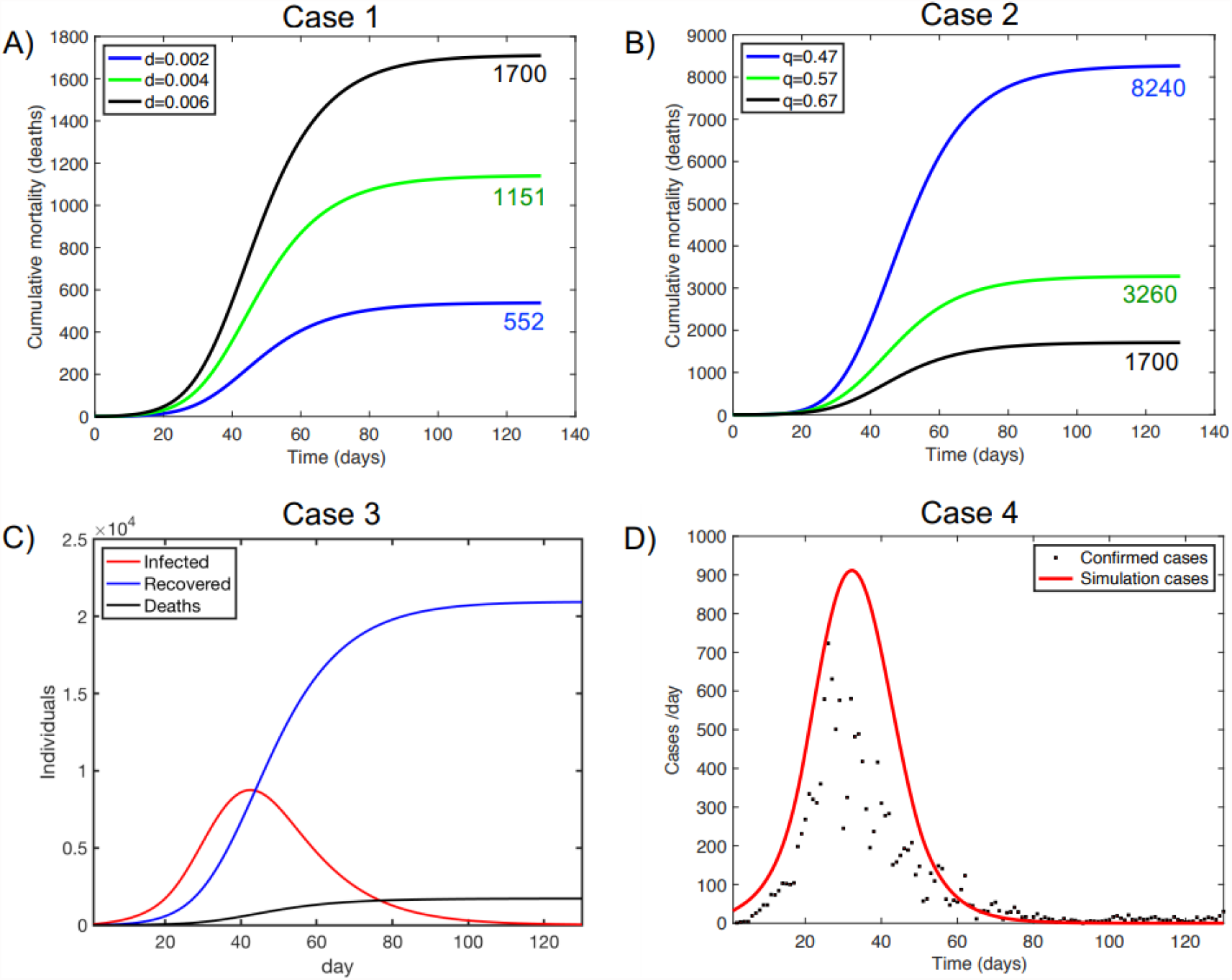
Model verification/evaluation against the first wave of the pandemic in the Basque Country. (A) Case 1: Simulation of cumulative mortality with varying disease mortality, (B) Case 2: Simulation of cumulative mortality with varying quarantine rate (day ^-1^), (C) Case 3: Simulation of infected, recovered and deaths (cumulative mortality), (D) Case 4: Model evaluation against new confirmed daily cases. Realistic initial conditions (Table 1) and parameter values (Table 2) were used. Simulation started on March 1 (day 1) and ended on July 8 (day 130).

**Figure 3.**
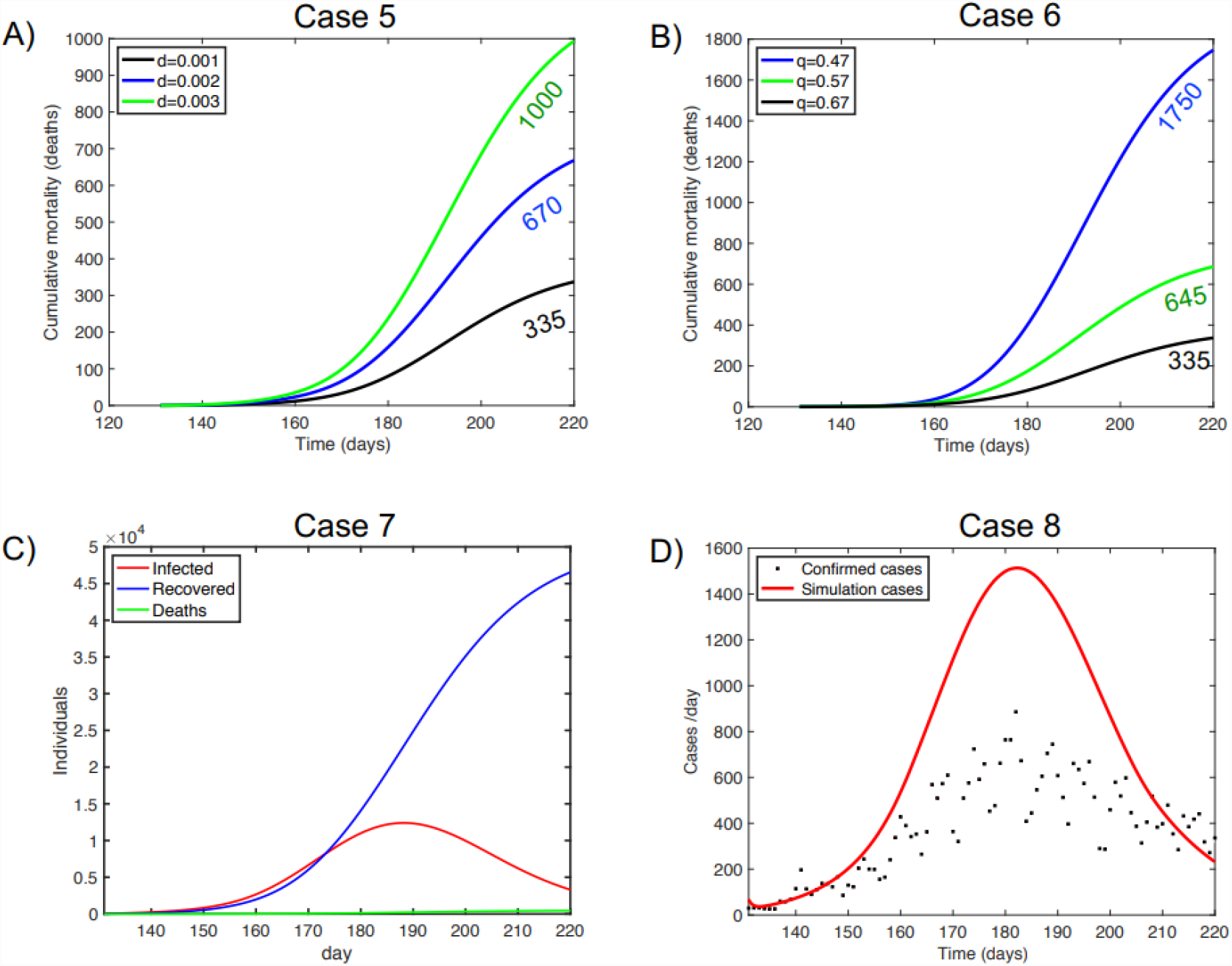
Model verification/evaluation against the second wave of the pandemic in the Basque Country (A) Case 5: Simulation of cumulative mortality with varying disease mortality, (B) Case 6: Simulation of cumulative mortality with varying quarantine rate (day ^-1^), (C) Case 7: Simulation of infected, recovered and deaths (cumulative mortality), (D) Case 8: Model evaluation against new confirmed daily cases. Realistic initial conditions (Table 1) and parameter values (Table 2) were used. The simulation started on July 9 (day 131) and ended on September 28 (day 220).

#### 2.9.1 First wave

Realistic scenarios with varying disease mortality (Figure 2A) and quarantine rate (Figure 2B) were simulated to verify and evaluate the performance of the model, focusing on the dynamics of the host-pathogen system in terms of cumulative mortality. In order to simulate the protection measure of the lockdown a tangential function was considered where the protection rate changed from a low protection level (5 ×10^−3^ day^-1^, before lockdown) to a high protection level (25 ×10^−3^ day^-1^, during lockdown) (see Table 2).

Increasing the disease mortality rate from low (2 ×10^−3^ day^-1^) to high (6 ×10^−3^ day^-1^) in Case 1 results in an expected increased cumulative mortality conforming to expectations (Figure 2A). For this simulation case, model evaluation against real cumulative data shows that the simulation with the highest disease mortality rate (6 ×10^−3^ day^-1^) results in 1700 deaths, showing the model has the best fit to the 1687 deaths confirmed by the BHD in the first wave (BHD, 2020).

Increasing the quarantine rate from low (5.7 ×10^−1^ day^-1^) to high (6.7 ×10^−1^ day^-1^) in Case 2 results in an expected reduced cumulative mortality (Figure 2B) conforming to model behaviour expectations. In this Case 2, model evaluation against real cumulative data shows the following: the simulation with the higher quarantine rate results in 1700 deaths, showing the model has the best fit to the 1687 deaths confirmed by the BHD in the first wave (BHD, 2020).

Once the model was verified and evaluated against mortality data, Case 3 was run to verify the model in terms of change of infected, recovered and death subpopulations with time during the first wave of the pandemic (Figure 2C). The behaviour of the model in these terms conforms to expectations regarding the number of recovered individuals (around 21000, approximately estimated cases – confirmed deaths) and fits to deaths (1687) confirmed by the BHD in the first wave of the pandemic (BHD, 2020).

Finally, Case 4 evaluated the model against new confirmed daily cases (Figure 2D). The model estimates 22230 cases as the number of true infections for the simulation period; about twice as high as the 13862 confirmed cases (BHD, 2020).

#### 2.9.2 Second wave

Similarly to the first wave, realistic scenarios with varying disease mortality (Figure 3A) and quarantine rate (Figure 2A) were simulated to verify and evaluate the performance of the model focusing on the dynamics of the host-pathogen system in terms of cumulative mortality. The simulations were run with specific initial conditions described in Table 1 and parameter values described in Table 2, except for (1) initial infected population of 40 individuals which was estimated from real data considering 33% of not confirmed infected people and (2) a low/medium-level protection rate of 6.5 × 10^−3^ day ^-1^; lower than in the first wave due to the end of lockdown, opening of indoors, mobility being allowed all over the country and no restrictions on international travellers entering the country.

In this second wave, simulation values for mortality rate were fitted using real deaths and quarantined individuals. These values were lower than those for the first wave since the disease incidence was much higher in young people, with a much lower fatality rate (BHD, 2020). Increasing disease mortality rate from low (1 × 10^−3^ day^-1^) to high (3 × 10^−3^ day^-1^) in Case 5 results in an expected increased cumulative mortality as expected (Figure 3A). For this case, model evaluation against real cumulative data shows that the simulation with the lower disease morality rate (1 × 10^−3^ day^-1^day^-1^) results in 335 deaths showing the model has the best fit to the 340 deaths confirmed by the BHD in the first wave (BHD, 2020).

Increasing the quarantine rate from low (5.7 × 10^−1^ day^-1^) to high (6.7 × 10^−1^ day^-1^) in Case 6 results in an expected reduced cumulative mortality (Figure 3B) as in Case 2, confirming the adequate behaviour of the model. In this Case 6, the simulation with the higher quarantine rate results in 335 deaths, showing the model has the best fit to the 340 deaths confirmed by the BHD in the first wave (BHD, 2020).

Case 7 verified the behaviour of the model in terms of changes in the infected, recovered and death subpopulations with time during the first wave of the pandemic (Figure 3C). The number of recovered individuals (around 46200) conforms to expectations considering the estimated cases and confirmed deaths, while estimated deaths (335) fits the number of fatalities confirmed by the BHD in the second wave of the pandemic (340) (BHD, 2020). Finally, in Case 8 the model estimated 45300 cases as the number of true infections for the simulation period (Figure 3D); higher than the 31000 confirmed cases (BHD, 2020).

### 2.10 Vaccination scenarios

Once the model was verified and evaluated, the impact of the vaccination strategy was tested considering five simulation scenarios for the fifth wave of the pandemic from March 7 to June 8, 2021. For these simulations, the varying non-pharmaceutical protection rates were from low-level to high-level protection rates as in Table 2: (i) the low-level protection rate α was estimated to be 5 × 10^−3^ day^-1^, including the use of masks, social distancing, opened indoor facilities, restaurants and bars, and easing of mobility limitations as they were initially relaxed inside the Basque territory and eventually all around Spain to promote tourism during the Easter holidays and before summer, (i) the medium-level protection rate of α=7.5 × 10^−3^ day^-1^ represented the previous scenario with no regional and national mobility restrictions, and (iii) the high-level protection rate of α=1 × 10^−2^ day^-1^ adds to the previous scenario the closure of indoor public and private facilities. In addition, in these simulations the transmission rate was increased to 1.15 day^-1^, since the Delta variant of the virus was known to spread significantly faster than the original version of the virus (Li et al., 2021). Initial conditions are those in Table 1 with changes in susceptible (S=2000000), infected cases (I=200) and vaccinated (V=52500) subpopulations due to the course of the disease dynamics.

#### 2.10.1 Simulation 1. Limited vaccine supply scenario: combination of Pfizer, Moderna, Astra Zeneca and Janssen

In this simulation, the model tries to mirror the vaccine strategy with a combination of Pfizer, Moderna, Astra Zeneca and Janssen vaccines followed by the BHD in the Basque Country. The vaccine-specific protection rates used in the model are those estimated in Table 4. The cumulative new daily confirmed cases for the simulation period were 48577 (black dots in Figure 4A). The model estimates 57000 cases and 55000 recovered individuals. Regarding fatalities, the simulation result (460 deaths) fits confirmed deaths (462) by the BHD for the simulation period (BHD, 2020).

**Figure 4.**
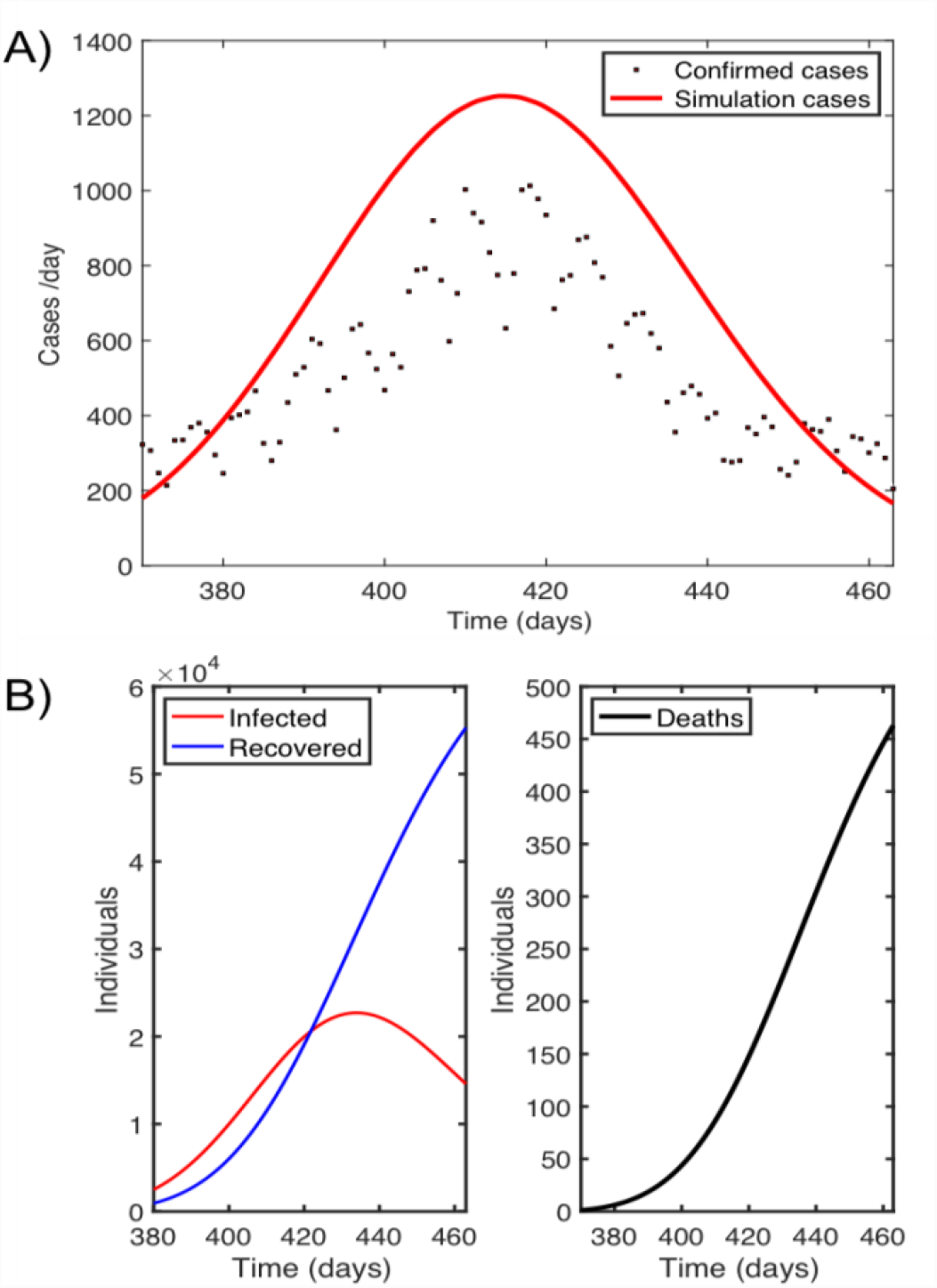
Limited vaccine supply scenario with a combination of Pfizer, Moderna, Astra Zeneca and Janssen vaccines: (A) Model evaluation against new confirmed daily cases. Realistic initial conditions (Table 1) and parameter values (Table 2) were used. The simulation started on March 7, 2021 (day 380) and ended on June 8, 2021 (day 460), (B) Simulation of infected, recovered and deaths (cumulative mortality), during vaccine administration.

For this vaccination scenario, simulations to test the effect of increasing the protection rate on new daily cases and cumulative mortality were run. The protection rate was increased from the adopted protection rate α=5 × 10^−3^ day^-1^ to α=1 × 10^−3^ day^-2^, in accordance with the strengthening of limitations in mobility and closure of indoor facilities such as restaurants and bars.

The highest number of new cases occurs with the lowest protection rate (α=5 × 10^−3^ day^-1^) where fewer restrictions are applied (Figure 5A). This number reduces to half when the protection measures increase to α= 7.5 × 10^−3^ day^-1^, decreasing even more if the protection rate is set to α=1 × 10^−2^ day^-1^. The same behaviour is observed in the cumulative mortality or death cases (Figure 5B).

**Figure 5.**
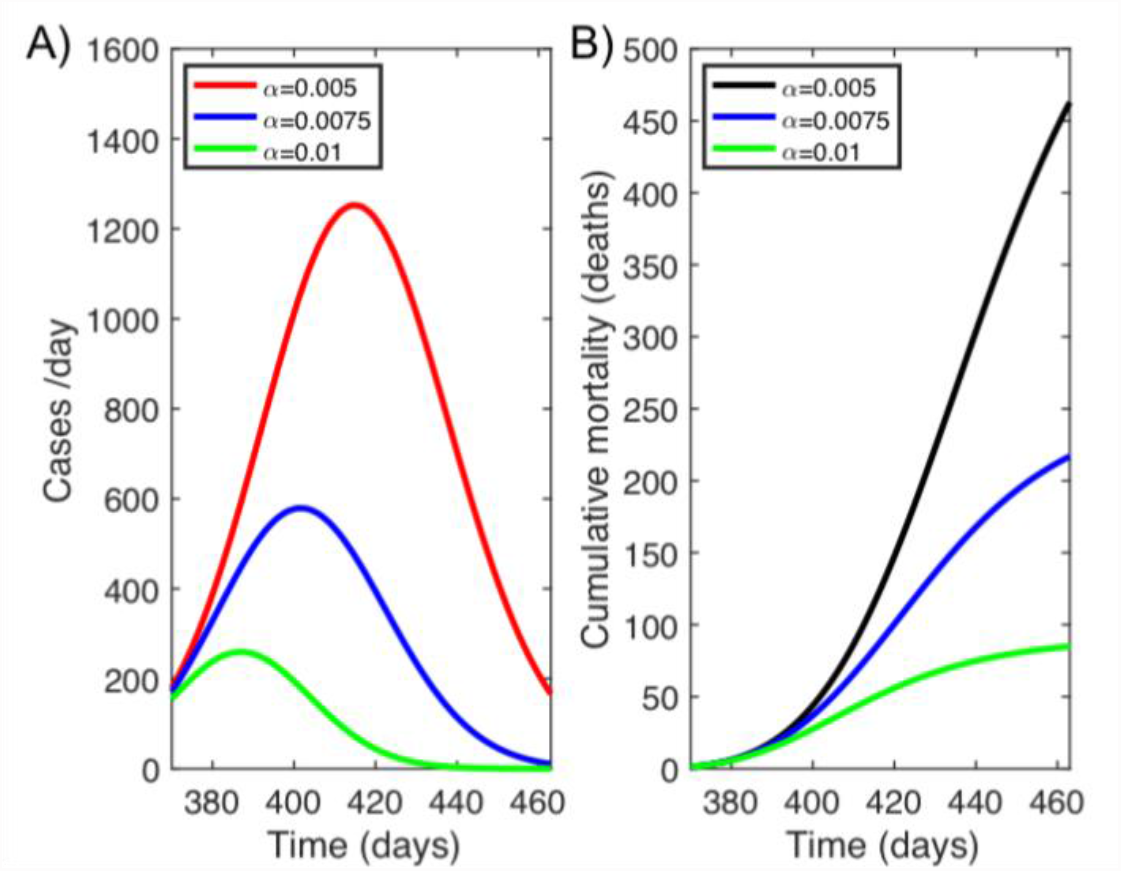
Simulations of new daily cases (A) and cumulative mortality rate (B) from March 7, 2021 (day 380) to June 8, 2021 (day 460) with varying protection rate (day^-1^) in a limited vaccine supply scenario with a combination of Pfizer, Moderna, Astra Zeneca and Janssen vaccines.

The following scenarios (3.3.2 – 3.3.5) are full supply scenarios where the type of vaccine can be chosen. Thus, the simulations contemplate the administration of a unique vaccine type, considering (i) the number of complete doses by June 8, the same as in the real limited vaccine supply scenario, and (ii) a vaccination rate that is the same for all vaccine types (see Table 4). For these vaccination scenarios, as in the first scenario, simulations to test the effect of increasing the protection rate (from α=0.004 to α=0.01) on new daily cases and cumulative mortality were run.

#### 2.10.2 Simulation 2. Pfizer scenario

In this simulation (Figure 6A), Pfizer is the unique vaccine administered to the population, with a vaccine protection rate of 1.7 × 10^−3^ day^-1^ (Table 4). The model estimates 51435 cumulative new cases and 415 deaths with the lowest protection rate; a lower number of fatalities compared to that obtained in the realistic limited vaccine supply scenario (462) (Simulation 1). Similarly, to simulation 1, an increased protection rate reduces the incidence of cases and mortality (Figure 6A), particularly when the protection rate is at its maximum with strong limitations in mobility and closure of indoor facilities such as restaurants and bars. In this protection scenario, the number of cases is about five times lower than that for a low protection rate, with no cases in the last 30 days. The cumulative mortality decreases from 415 to 85 deaths.

**Figure 6.**
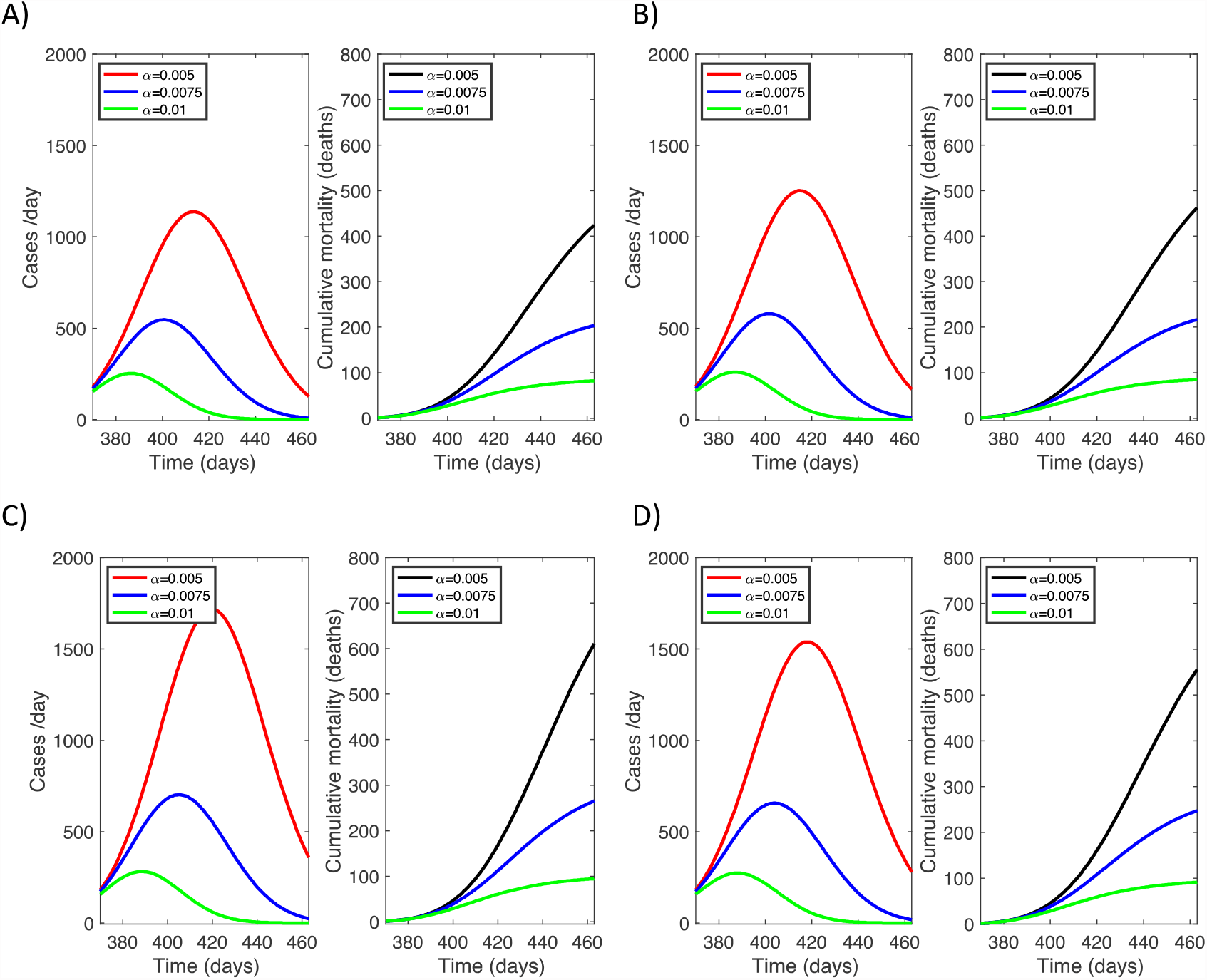
Simulation of new daily cases and cumulative mortality (deaths) with a varying protection rate in a unique vaccine administration scenario: A) Pfizer, B) Moderna, C) Astra Zeneca and D) Janssen.

#### 2.10.3 Simulation 3. Moderna scenario

In this simulation (Figure 6B), Moderna is the unique vaccine administered to the population with a vaccine protection rate of 1.6 × 10^−3^ day^-1^ (Table 4). The model estimates 56,500 cases in the simulation period and 458 deaths with the lowest protection rate. The impact of the high protection scenario (green line) on disease dynamics is also high; cumulative mortality decreases from 458 to 87 deaths.

#### 2.10.4 Simulation 4. Astra Zeneca scenario

This simulation represents a scenario with the Astra Zeneca vaccine as the unique vaccine administered (Figure 6C), with a vaccine protection rate of 1 × 10^−3^ day ^-1^ (Table 4). The model in this case estimates 77,900 cases in the simulation period and 600 deaths with the lowest protection rate. Here, for the medium protection level the model estimates 265 deaths. However, for the higher protection rate, differences between the responses to vaccines are not significant.

#### 2.10.5 Simulation 5. Janssen scenario

This simulation (Figure 6D) shows the Janssen vaccine as the unique vaccine administered with a vaccine protection rate of 1.1 × 10^−3^ day −1(Table 4). For the highest protection rate, new daily cases and cumulative mortality, are similar to those observed for the other vaccines. Nonetheless, for the medium and, particularly, for the lowest protection rate, the Janssen vaccine estimates 62,030 cases in the simulation period and 550 deaths.

## 3 Discussion and conclusions

This contribution covers the theoretical and mathematical basis for modelling dynamics and epidemiology of COVID-19, specifically focusing on the effect of the interaction between the initial phase of the vaccination and non-pharmaceutical protection measures such as self-protection, mobility restrictions, closure of indoor facilities and lockdown. The Kermack and McKendrick (1927) epidemiological theory was adapted to build a SEIR deterministic model for COVID-19 to assess the impact of this interaction on infection cases and disease mortality. The model was verified and validated using the response timeline, vaccination strategies and non-pharmaceutical interventions implemented in the Basque Country (N Spain). Although robust validation of the model predictions is needed, initial results and evaluation show the potential of the model to be easily modified to match other regions’ or countries’ timelines, or the different response strategies implemented in other countries.

The waves of the epidemic curve (confirmed cases) in this region can be discussed in terms of the non-pharmaceutical interventions and disease management. Fifteen days after the first case was confirmed in the Basque Country, schools were closed in all regions of Spain and a nationwide lockdown (confinement) was declared, banning all public events. In this context, during the first wave Covid-19 testing management and coverage was limited. Tests were done only, for those who presented symptoms such as fever and cough. People who did not seek medical attention were tested very rarely. The second wave peak was reached in late summer when mobility all around the country was permitted, international travellers entered the country without restrictions, and indoor public and private facilities were open. The peak of the third wave reached on November linked with the return to schools and work, and the increase of indoor activities. A **fourth wave** peak was recorded on January 2021 linked with Christmas holidays despite the limitations imposed by the government, limiting gatherings of people and mobility in the territory. The **fifth wave** was linked with the easing of mobility limitations inside the Basque territory to promote tourism during the Easter holidays.

The projections explored here for model verification and validation do not differ much from the reported cumulative mortality data and are consistent with the descriptive analysis of the epidemic curve and disease dynamics found by other similar modelling studies in the Basque Country (López & Rodó, 2020). However, estimated new daily cases are significantly higher than those reported by the BHD (BHD, 2020). This result is consistent with the fact that confirmed cases may be undercounted (Giattino, 2020) since one of the key limitations when modelling this disease is that the reported cases only become confirmed cases by a test, and there are a substantial proportion of infected people that never get tested, particularly in the first wave, because they were asymptomatic or never sought medical assistance. The model here, as well as others of this type or more sophisticated ones, use confirmed cases and deaths, testing rates, and a range of assumptions and epidemiological knowledge to estimate this proportion and consequently show a higher number of cases compared to the reported data (Giattino, 2020). This is also consistent with the seroprevalence study carried out in Spain, where around 33% of the infected cases were asymptomatic (Pollán et al., 2020). On the other hand, there is a deviation ratio and as expected if the number of cases increase the range between model prediction and real data increases.

The simulation explored here for the limited vaccine supply scenario confirms that the model is correctly validated against the real data. The performance when increasing the protection measures from low-level to high-level non-pharmaceutical protection measures, follows the expected decreasing trend of COVID-19 cases and cumulative mortality. Comparing this scenario to the full vaccine supply scenarios, results suggest that the ideal scenario for limiting the impact of OCVID-19 is the one combining vaccination and high protection levels for non-pharmaceutical measures. There is not a big variation between vaccines in terms of cases and cumulative mortality when the protection rate is high (Figures 5 and 6). Differences in disease incidence response between vaccines need to be taken with caution since there may be some unpredictable latent covariates as described in section 2.7. asaas described in section *2*.*7 Model limitations*.

Overall, the results suggest that in an initial vaccination phase (30%-50% of the population is vaccinated) COVID-19 incidence, as measured on daily cases and cumulative mortality, importantly decreases when vaccination and a high level of non-pharmaceutical interventions are in place. That is, in the first vaccination phase, together with vaccination, strong mobility restrictions and closure of indoor facilities such as public spaces, restaurants and bars are critical to significantly control disease outbreaks. When the adopted measures are in the low level (no mobility restrictions and indoor facilities open), COVID-19 cases and deaths remain too high to contain the outbreak. As a positive result, it seems that the number of fatalities has decreased with respect to previous waves. This may be explained by the vaccine programme, which prioritises the most vulnerable people by age (BHD, 2020). However, cases are still high, thus, regarding COVID-19 cases, it bears repeating that model results also suggest that initial phase vaccination may synergise with other non-pharmaceutical measures, until the proportion of the immune population increases.

The relevance of these results lies in the fact that they support research regarding the relative importance of the non-pharmaceutical measures and vaccination involved in the termination of the epidemic. One limitation of the studied model approach is the assumption that most parameters, except the protection measure in the first wave, take fixed values independent of time. The assumption of constancy in time has the advantage of simplifying the models and facilitates its use. However, both the prevalence of infection and the transmission of the virus may be tied to environmental conditions (Eslami & Jalili, 2020). For the sake of simplicity and the obtention of a preliminary picture, in this model, population has not been divided by age groups. This age-structure should be considered to improve the present model as the vaccination programme has been prioritised by age groups. An age-structured version of this model would give a more accurate picture of the virus transmission (Foy et al., 2021), same as considering the new variants of the virus that could directly impact in the vaccine induced protection and the transmission rate (Moore, 2021).

Given the current pandemic caused by the transmission of SARS-CoV-2, the construction of mathematical models such as this one, based on epidemiological data, has allowed us to describe the interactions, explain the dynamics of infection, as well as predict possible scenarios that may arise with the introduction of measures such as social distancing, the use of masks, mobility limitations and vaccination programmes. Mathematical models are highly relevant for making objective and effective decisions to control the disease. These models have supported and will continue to contribute to the selection and implementation of programmes and public policies that prevent associated complications, slow down the spread of the virus and minimise the appearance of severe cases of disease that may collapse health systems.

## Data Availability

All data produced in the present work are contained in the manuscript

## 4 Acknowledgments

This investigation was conducted under the framework of the Master in Public Health of the University of Basque Country (UPV/EHU) (Department of Preventive Medicine and Public Health). We appreciate this support. The COVID-19 incidence and mortality data were obtained from the Basque Health Department (BHD) (Osakidetza) (Open Data Euskadi web page: https://opendata.euskadi.eus/catalogo/-/evolucion-del-coronavirus-covid-19-en-euskadi/). The contents of this manuscript do not necessarily reflect the point of view of the BHD and in no ways anticipate the BHD’s future policy in this area. The manuscript benefited from helpful discussions with Dr. Aitana Lertxundi and Dr. Naroa Kajarabille (Department of Preventive Medicine and Public Health, UPV/EHU) and Ane Murueta (Department of Neurosciences, UPV/EHU), and examination of the model structure by Dr. Tal Ben-Horin (Department of Clinical Sciences, North Carolina State University) and Morganne Igoe (Department of Mathematics, University of Tennessee).

## Notes

### Competing Interest Statement

The authors have declared no competing interest.

### Funding Statement

This study did not receive any funding

### Author Declarations

This study involves only openly available human data, which can be obtained from Open Data Euskadi web page: https://opendata.euskadi.eus/catalogo/-/evolucion-del-coronavirus-covid-19-en-euskadi/).

### Summary of Updates

This new version includes a section for model limitations. The equation system has been corrected.

